# A randomized, double-blind phase I clinical trial of two recombinant dimeric RBD COVID-19 vaccine candidates: safety, reactogenicity and immunogenicity

**DOI:** 10.1101/2021.10.04.21264522

**Authors:** Sonia Pérez-Rodríguez, Meiby de la Caridad Rodríguez-González, Rolando Ochoa-Azze, Yanet Climent-Ruiz, Carlos Alberto González-Delgado, Beatriz Paredes-Moreno, Carmen Valenzuela-Silva, Laura Rodríguez-Noda, Rocmira Perez-Nicado, Raúl González-Mugica, Marisel Martínez-Pérez, Belinda Sánchez-Ramírez, Tays Hernández-García, Alina Díaz-Machado, Maura Tamayo-Rodríguez, Alis Martín-Trujillo, Jorman Rubino-Moreno, Anamary Suárez-Batista, Marta Dubed-Echevarría, María Teresa Pérez-Guevara, Mayté Amoroto-Roig, Yanet Chappi-Estévez, Gretchen Bergado-Báez, Franciscary Pi-Estopiñán, Guang-Wu Chen, Yuri Valdés-Balbín, Dagmar García-Rivera, Vicente Vérez-Bencomo

**Author notes:** **Corresponding author:** Rolando Ochoa-Azze, MD, PhD. Finlay Vaccine Institute. 21^st^ Ave. Nº 19810 between 198 and 200 Streets, Atabey, Playa, Havana, Cuba. Joint first authors (Sonia Pérez-Rodríguez, Meiby de la Caridad Rodríguez-González, Rolando Ochoa-Azze).

## Abstract

**Background:** The Receptor Binding Domain (RBD) of the SARS-CoV-2 spike protein is the target for many COVID-19 vaccines. Here we report results for phase 1 clinical trial of two COVID-19 vaccine candidates based on recombinant dimeric RBD (d-RBD).

**Methods:** We performed a randomized, double-blind, phase I clinical trial in the National Centre of Toxicology in Havana. Sixty Cuban volunteers aged 19-59 years were randomized into three groups (20 subjects each): 1) FINLAY-FR-1 (50 mcg d-RBD plus outer membrane vesicles from *N. meningitidis*); 2) FINLAY-FR-1A-50 mcg d-RBD (three doses); 3) FINLAY-FR-1A-25 mcg d-RDB (three doses). The FINLAY-FR-1 group was randomly divided to receive a third dose of the same vaccine candidate (homologous schedule) or of FINLAY-FR-1A-50 (heterologous schedule). The primary outcomes were safety and reactogenicity. The secondary outcome was vaccine immunogenicity. Humoral response at baseline and following each vaccination was evaluated using live-virus neutralization test, anti-RBD IgG ELISA and *in-vitro* neutralization test of RBD:hACE2 interaction.

**Results:** Most adverse events were of mild intensity (63.5%), solicited (58.8%), and local (61.8%); 69.4% with causal association with vaccination. Serious adverse events were not found. The FINLAY-FR-1 group reported more adverse events than the other two groups. After the third dose, anti-RBD seroconversion was 100%, 94.4% and 90% for the FINLAY-FR-1, FINLAY-FR-1A-50 and FINLAY-FR-1A-25 respectively. The *in-vitro* inhibition of RBD:hACE2 interaction increased after the second dose in all formulations. The geometric mean neutralizing titres after the third dose rose significantly in the group vaccinated with FINLAY-FR-1 with respect to the other formulations and the COVID-19 Convalescent Serum Panel. No differences were found between FINLAY-FR-1 homologous or heterologous schedules.

**Conclusions:** Vaccine candidates were safe and immunogenic, and induced live-virus neutralizing antibodies against SARS-CoV-2. The highest values were obtained when outer membrane vesicles were used as adjuvant.

**Trial registry:** https://rpcec.sld.cu/en/trials/RPCEC00000338-En

## 1. Introduction

The COVID-19 pandemic persists, with high incidence and mortality rates [1]. COVID-19 is caused by SARS-CoV-2, an enveloped positive-sense RNA virus, with four main structural proteins: spike (S, on its surface), membrane, envelope, and nucleocapsid proteins [2,3].

The trimeric S glycoprotein mediates the attachment to the human angiotensin-converting enzyme (hACE2) on host cells surface. S protein has two subunits: S1 and S2. S1 mediates hACE2 binding through the receptor binding domain (RBD), while S2 mediates viral fusion [2,3].

Neutralizing antibodies against SARS-CoV-2 are mainly stimulated by the RBD, while other SARS-CoV-2 proteins can promote an immunopathogenic mechanism mediated by antibodies known as ADE (Antibody Dependent Enhancement) [4-6]. Blocking RBD-hACE2 interaction is the main target for vaccines against SARS-CoV-2 [6-8].

Several vaccines have been developed, based on different platforms. The World Health Organization (WHO) has approved the use of inactivated virus vaccines, adenovirus vector vaccines and mRNA vaccines, which have demonstrated their efficacy against COVID-19, especially those based on new technologies; however, they have raised safety concerns [7,9,10]. Another approach has been the development of protein subunit vaccines, especially those using the RBD of the spike protein of SARS-CoV-2 [10-12].

FINLAY-FR-1 (SOBERANA 01) and FINLAY-FR-1A (SOBERANA Plus) are recombinant dimeric RBD (d-RBD) vaccine candidates. They are produced under Good Manufacturing Practice conditions at The Finlay Vaccine Institute and The Centre of Molecular Immunology, in Havana, Cuba; both finished the preclinical and toxicological evaluations.

FINLAY-FR-1A and FINLAY-FR-1 are adsorbed on alum; FINLAY-FR-1 also has outer membrane vesicles from *Neisseria meningitidis* group B (OMVs) as adjuvant. Its adjuvant role has been well-documented, inducing a strong immune response and a Th1 pattern [13-15].

This study evaluated and compared safety and reactogenicity of FINLAY-FR-1A and FINLAY-FR-1, and explored the immunogenicity induced by three doses of these vaccine candidates.

## 2. Methods

### 2.1. Study design and participants

This phase I, randomized, double-blind clinical trial was carried out at the National Centre of Toxicology (CENATOX) in Havana, Cuba. Sixty Cuban volunteers aged 19-59 years, with body mass index 18.5-29.9 kg/m^2^, of both sexes, were recruited (Table 1). Two vaccine candidates were evaluated: FINLAY-FR-1 and FINLAY-FR-1A, this one at two d-RBD concentrations: 25 mcg (FINLAY-FR-1A-25) and 50 mcg (FINLAY-FR-1A-50). Participants were distributed into three 20 subjects groups: FINLAY-FR-1; FINLAY-FR-1A-50 and FINLAY-FR-1A-25.

**Table 1:**
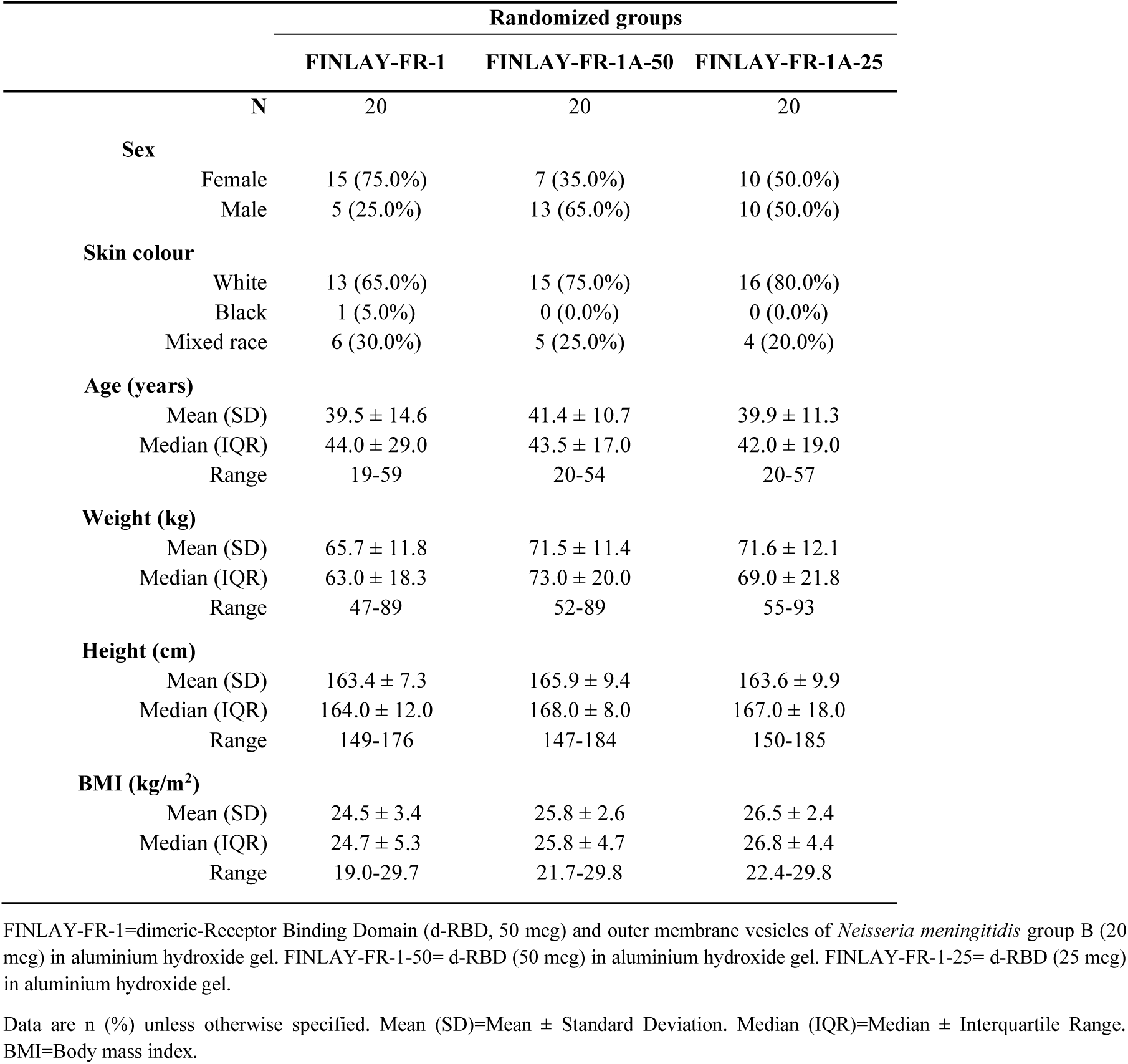
Baseline demographic characteristics of subjects included in the clinical trial.

|  |  | Randomized groups |  |  |
| --- | --- | --- | --- | --- |
|  |  | FINLAY-FR-1 | FINLAY-FR-1A-50 | FINLAY-FR-1A-25 |
| N |  | 20 | 20 | 20 |
| **Sex** |  |  |  |  |
|  | Female | 15 (75.0%) | 7 (35.0%) | 10 (50.0%) |
|  | Male | 5 (25.0%) | 13 (65.0%) | 10 (50.0%) |
| **Skin colour** |  |  |  |  |
|  | White | 13 (65.0%) | 15 (75.0%) | 16 (80.0%) |
|  | Black | 1 (5.0%) | 0 (0.0%) | 0 (0.0%) |
|  | Mixed race | 6 (30.0%) | 5 (25.0%) | 4 (20.0%) |
| **Age (years)** |  |  |  |  |
|  | Mean (SD) | 39.5 ± 14.6 | 41.4 ± 10.7 | 39.9 ± 11.3 |
|  | Median (IQR) | 44.0 ± 29.0 | 43.5 ± 17.0 | 42.0 ± 19.0 |
|  | Range | 19-59 | 20-54 | 20-57 |
| **Weight (kg)** |  |  |  |  |
|  | Mean (SD) | 65.7 ± 11.8 | 71.5 ± 11.4 | 71.6 ± 12.1 |
|  | Median (IQR) | 63.0 ± 18.3 | 73.0 ± 20.0 | 69.0 ± 21.8 |
|  | Range | 47-89 | 52-89 | 55-93 |
| **Height (cm)** |  |  |  |  |
|  | Mean (SD) | 163.4 ± 7.3 | 165.9 ± 9.4 | 163.6 ± 9.9 |
|  | Median (IQR) | 164.0 ± 12.0 | 168.0 ± 8.0 | 167.0 ± 18.0 |
|  | Range | 149-176 | 147-184 | 150-185 |
| **BMI (kg/m<sup>2</sup>)** |  |  |  |  |
|  | Mean (SD) | 24.5 ± 3.4 | 25.8 ± 2.6 | 26.5 ± 2.4 |
|  | Median (IQR) | 24.7 ± 5.3 | 25.8 ± 4.7 | 26.8 ± 4.4 |
|  | Range | 19.0-29.7 | 21.7-29.8 | 22.4-29.8 |

FINLAY-FR-1=dimeric-Receptor Binding Domain (d-RBD, 50 mcg) and outer membrane vesicles of *Neisseria meningitidis* group B (20 mcg) in aluminium hydroxide gel. FINLAY-FR-1-50= d-RBD (50 mcg) in aluminium hydroxide gel. FINLAY-FR-1-25= d-RBD (25 mcg) in aluminium hydroxide gel.
Data are n (%) unless otherwise specified. Mean (SD)=Mean ± Standard Deviation. Median (IQR)=Median ± Interquartile Range. BMI=Body mass index.

All participants underwent a screening visit (full medical history, pregnancy rapid test in women of childbearing potential, SARS-CoV-2 PCR tests, blood tests (HIV; hepatitis B and C serology; full blood count; kidney and liver function tests; background of IgG anti-SARS-CoV-2 antibodies, and virus neutralization test). Exclusion criteria were: history of COVID-19, SARS-CoV-2 PCR-positive tests or detection of antibodies anti-SARS-CoV-2, any severe disease or decompensated chronic disease, immunodeficiency, history of serious allergy, pregnancy, breastfeeding, and immunological treatment during the last 30 days.

The study was registered at the Cuban Public Registry of Clinical Trials: https://rpcec.sld.cu/en/trials/RPCEC00000338-En, included in WHO International Clinical Registry Trials Platform.

### 2.2. Ethical considerations

The Cuban Ministry of Public Health (MINSAP), the Independent Ethics Committee (IEC) for Studies on Human Subjects, at CENATOX and the Cuban National Regulatory Agency (Centre for State Control of Medicines and Medical Devices, CECMED), approved the trial and the procedures (CECMED, Authorization date: 13/10/2020, Reference number: 05.013.20BA). It was conducted according to the Declaration of Helsinki and Good Clinical Practice.

An Independent Data Monitoring Committee (IDMC) analysed safety, reactogenicity, and immunogenicity data. During recruitment, investigators provided potential participants with extensive oral and written information. The decision to participate in the study was completely voluntary. Written informed consent was obtained from all volunteers. During the study, the IEC and IDMC assessed the trial’s risk-benefit ratio and assured the rights, health and privacy of volunteers, including information confidentiality.

### 2.3. Product under evaluation

Vaccine antigen: SARS-CoV-2 RBD (sequence: 319-541 amino acid residues with a poly-histidine fusion tag at its C-terminus), expressed in CHO cells. RBD is dimerized through a Cys538-Cys538 interchain disulphide bridge.

FINLAY-FR-1A (SOBERANA Plus) vaccine candidate was evaluated at two d-RBD concentrations. Composition per dose (0.5 mL): d-RBD 50 mcg or 25 mcg, NaCl 4.250 mg, Na_2_HPO4·0.03 mg, NaH_2_PO4·0.02 mg, thiomersal 0.05 mg, injection water, aluminium hydroxide gel 1.25 mg, pH 6.0-7.2.

FINLAY-FR-1 (SOBERANA 01) vaccine candidate composition: d-RBD 50 mcg, OMVs 20 mcg, NaCl 4.250 mg, Na_2_HPO4·0.03 mg, NaH_2_PO4·0.02 mg, thiomersal 0.05 mg, injection water, aluminium hydroxide gel 1.25 mg, pH 6.0–7.2.

### 2.4. Randomization and blinding

Stratified random blinded sampling was used to select the sample of the universe of Cuban citizens aged 19-59 years, which was proportionally divided in two age subgroups: 19-39 and 40-59 years to ensure a proper representation of each age subgroup. Allocation of participants in each vaccine group was done by simple random blinded sampling using a centralized technology. Each participant got an identification code, which matched the vaccine vial label code.

All study staff, investigators, sponsor personnel and subjects, remained blinded until the conclusion of the study (28 days after the last dose of the vaccine was applied to all volunteers). All vials had the same characteristics: R2 vial, single dose, volume and pink cap.

### 2.5. Procedures

After medical screening, 60 eligible participants were randomly allocated to three groups: FINLAY-FR-1, FINLAY-FR-1A-50 and FINLAY-FR-1A-25 (Figure 1). All participants received three vaccine doses: FINLAY-FR-1 and FINLAY-FR-1A-50 groups were vaccinated on days T0 (initial); second, on day 28 (T28); and third, between 65 and 73 days after the second dose. For the third dose, the FINLAY-FR-1 group was randomly divided in two 10 participants subgroups: one received FINLAY-FR-1 (homologous schedule); the other subgroup received FINLAY-FR-1A-50 (heterologous schedule). The FINLAY-FR-1A-25 group was vaccinated with three doses: T0, T28 and T56.

**Fig.1.**
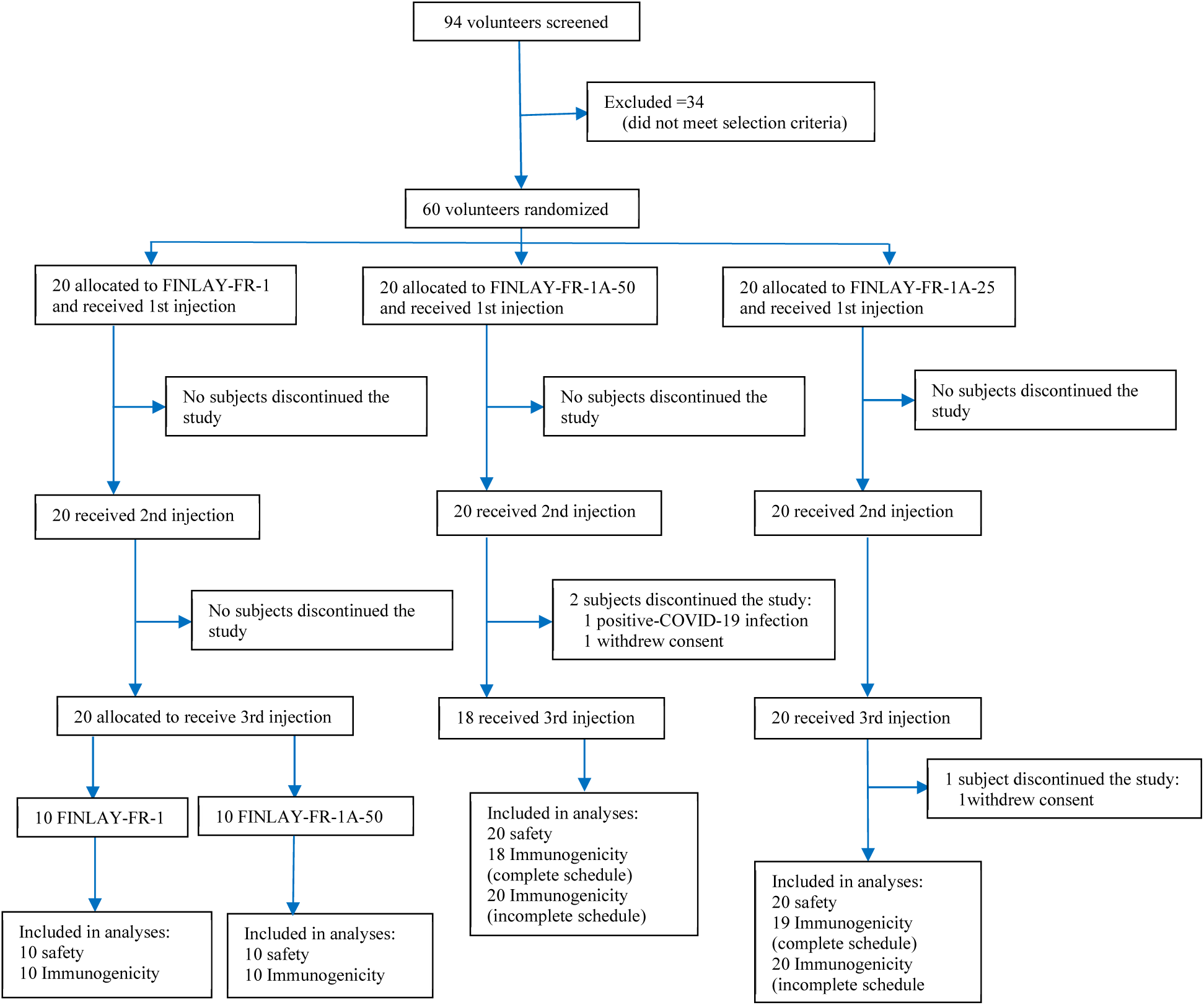
Disposition of subjects. Trial profile. FINLAY-FR-1=dimeric-Receptor Binding Domain (d-RBD, 50 mcg) and outer membrane vesicles of *Neisseria meningitidis* group B (20 mcg) in aluminium hydroxide gel. FINLAY-FR-1-50=d-RBD (50 mcg) in aluminium hydroxide gel. FINLAY-FR-1-25=d-RBD (25 mcg) in aluminium hydroxide gel.

For immunological tests, blood samples were collected on days 0 (before vaccination), T28, T56, and 28 days after the third dose. For haematology and blood chemistry tests, blood samples were collected before vaccination, and 28 days after the last dose.

Volunteers were closely observed for 3 h post-vaccination. After each dose, active surveillance was carried out on days 1 (vaccination), 2, 3, and 7 for all groups. For passive surveillance, participants were instructed to complete a diary record of solicited local and systemic adverse reactions during the follow-up period.

Solicited and protocol-defined local site reactions (injection site pain, redness, warmth, swelling, induration) and systemic symptoms (general malaise, rash, and fever defined as an axillary temperature ≥38°C) were recorded for 7 days after each dose. All other events were recorded throughout the follow-up period. The severity of expected and protocol-defined local and systemic adverse events were graded as mild, moderate and severe, according to Brighton Collaboration definition and the Common Terminology Criteria for Adverse Events version 5·0. Severity of unsolicited adverse events were graded as: mild (transient or mild discomfort, no interference with activity), moderate (mild to moderate limitation in activity), severe (marked limitation in activity) [16,17].

All adverse events were reviewed for causality, and events were classified according to WHO: inconsistent causal association to immunization, consistent causal association to immunization, indeterminate, or unclassifiable [18].

#### 2.5.1. Humoral immune response at baseline and following vaccination was evaluated by

a. *in-house indirect quantitative IgG anti-RBD ELISA*, using d-RBD as coating antigen and an anti-human-γ:peroxidase conjugate. This assay uses an in-house standard characterized serum, which was arbitrarily assigned 200 AU/mL. The standard curve was constructed by performing six two-fold serial dilutions: from 1:100 to 1:1600. Serum samples were diluted from 1:100 to 1:400. The IgG anti-RBD concentration was determined by interpolating the optical density of serum samples in the standard curve constructed using four-parameter log-logistic function [19].
b. *Molecular virus neutralization test*, based on antibody-mediated blockage of RBD:hACE2 interaction. This test is an in-vitro surrogate of the live-virus neutralization test. It uses recombinant RBD-mouse-Fc (RBD-Fcm) and the host cell receptor hACE2-Fc (ACE2-Fch) as coating antigen. Human antibodies against RBD can block the interaction of RBD-Fcm with ACE2-Fch. The RBD-Fcm that was not inhibited can bind to ACE2-Fch, and it is recognized by a monoclonal antibody anti-γ murine conjugated to alkaline phosphatase. The inhibition ratio of RBD:hACE2 interaction at a serum dilution of 1:100 and the half-maximal molecular virus neutralization titres (mVNT_50_) were calculated [20].
c. *Conventional live-virus neutralization test*. This is the gold standard for determining antibody efficacy against SARS-CoV-2. It is a colorimetric assay based on antibody neutralization of SARS-CoV-2 live virus cytophatic effect on Vero E6 cells. It was used the D614G variant that was circulating. The conventional live-virus neutralization titres (cVNT) were calculated [21].

The vaccine-elicited humoral immune response was compared with that of the Cuban Convalescent Serum Panel (CCSP), composed of 68 serum samples from asymptomatic individuals (25), and those recovered from mild/moderate (30) and serious (13) COVID-19. This panel was previously characterized by standardized ELISA, in-vitro inhibitory assay and live-virus neutralization test.

### 2.6. Outcomes

The two co-primary outcomes, safety and reactogenicity, were assessed until 28 days after the third, last dose. Safety was measured by the occurrence of serious adverse events. Results of laboratory analyses on blood samples at 28 days after the last dose were compared to pre-vaccination values. The secondary outcome, vaccine immunogenicity, was estimated after vaccination, as explained in “Procedures”, and compared to baseline.

### 2.7. Statistical analysis

Calculation of the sample size was based on a serious adverse events rate lower than 5%. Two-sided 95% confidence intervals for one proportion were calculated, taking into account a target width of 0.25. Safety and reactogenicity endpoints were described as frequencies (%). The following values were reported: mean, standard deviation (SD), median, interquartile range, and range, for the demographic characteristics and adverse events; median, for immunological endpoints; geometric mean titres (GMT) and 95% confidence intervals (CI), for mVNT_50_ and cVNT. Seroconversion rates for IgG anti-RBD antibodies (≥4-fold increase in antibody titres over pre-immunization titres) were calculated for each subject.

The Student’s t-Test, the Wilcoxon Signed-Rank Test or Kruskal Wallis Test were used for before-after statistical comparison. Statistical analyses were done using SPSS version 25·0; EPIDAT version 4.1, Prism GraphPad version 6.0. A type I error of 0·05 was used.

## 3. Results

From October 19, 2020, to October 24, 2020, 94 volunteers were enrolled into the study; 34 participants were excluded for not meeting selection criteria and 60 volunteers were randomized into the three experimental groups. There were three treatment interruptions: one due to COVID-19 infection after the second dose with FINLAY-FR-1A-50 and two voluntary dropouts: one subject in that same group and another in the FINLAY-FR-1A-25.

All randomized subjects were included in the safety analysis. Immunogenicity with the 3-dose schedule was evaluated in most subjects, except those with treatment interruptions after the second dose (Figure 1).

The demographic characteristics are summarized in Table 1. The groups were homogeneous for all the variables studied, except sex, with a predominance of women in the FINLAY-FR-1 group, and men in the FINLAY-FR-1A-50 group. White skin colour predominated and the median age was around 43 years (Table 1).

An adverse event was reported by 80% of participants. There were 170 adverse events of 31 different types. Most adverse events were of mild intensity (63.5%), solicited (58.8%), and local (61.8%); 69.4% had a causal association with vaccination, predominating pain at the vaccination site (Table 2 and 3). Most adverse events appeared in the first 24 hours and 59.6% lasted less than 24 hours. No serious adverse events were found, only 4 (2.4%) were severe.

**Table 2:** Main characteristics of adverse events following vaccination with d-RBD vaccine candidates.

|  | Vaccine Candidates |  |  | Total |
| --- | --- | --- | --- | --- |
|  | FINLAY-FR-1 | FINLAY-FR-1A-50 | FINLAY-FR-1A-25 |  |
| N | 20 | 20 | 20 | 60 |
| Subjects with some AE | 18 (90.0%) | 17 (85.0%) | 13 (65.0%) | 48 (80.0%) |
| Subjects with some VAAE | 17 (85.0%) | 15 (75.0%) | 12 (60.0%) | 44 (73.3%) |
| Subjects with some Serious AE | 0 (0.0%) | 0 (0.0%) | 0 (0.0%) | 0 (0.0%) |
| Subjects with some Serious VAAE | 0 (0.0%) | 0 (0.0%) | 0 (0.0%) | 0 (0.0%) |
| Subjects with some Severe AE | 1 (5.0%) | 2 (10.0%) | 3 (15.0%) | 6 (10.0%) |
| Subjects with some Severe VAAE | 1 (5.0%) | 2 (10.0%) | 1 (5.0%) | 4 (6.7%) |
| **Total Adverse Events** | **83** | **52** | **35** | **170** |
| Mild AE | 49 (59.0%) | 42 (80.8%) | 17 (48.6%) | 108 (63.5%) |
| Moderate AE | 33 (39.8%) | 8 (15.4%) | 15 (42.9%) | 56 (32.9%) |
| Severe AE | 1 (1.2%) | 2 (3.8%) | 3 (8.6%) | 6 (3.5%) |
| Serious AE | 0 (0.0%) | 0 (0.0%) | 0 (0.0%) | 0 (0.0%) |
| Local AE | 52 (62.7%) | 29 (55.8%) | 24 (68.6%) | 105 (61.8%) |
| Systemic AE | 31 (37.3%) | 23 (44.2%) | 11 (31.4%) | 65 (38.25) |
| VAAE | 58 (69.9%) | 34 (65.4%) | 26 (74.3%) | 118 (69.4%) |
| Serious VAAE | 0 (0.0%) | 0 (0.0%) | 0 (0.0%) | 0 (0.0%) |
| Severe VAAE | 1 (1.2%) | 2 (3.8%) | 1 (2.9%) | 4 (2.4%) |
| Reported Severe VAAE | Pain | Pain/Redness | Swelling |  |

FINLAY-FR-1=dimeric-Receptor Binding Domain (d-RBD, 50 mcg) and outer membrane vesicles of *Neisseria meningitidis* group B (20 mcg) in aluminium hydroxide gel. FINLAY-FR-1-50= d-RBD (50 mcg) in aluminium hydroxide gel. FINLAY-FR-1-25= d-RBD (25 mcg) in aluminium hydroxide gel.
Data are n (%). AE=Adverse Event. VAAE=Vaccine-Associated Adverse Event.

**Table 3:** Frequency of vaccine-associated adverse events following vaccination with d-RBD vaccine candidates.

|  | Vaccine Candidates |  |  | Total |
| --- | --- | --- | --- | --- |
|  | FINLAY-FR-1 | FINLAY-FR-1A-50 | FINLAY-FR-1A-25 |  |
| N | 20 | 20 | 20 | 60 |
| **Subjects with some VAAE** | 17 (85.0%) | 15 (75.0%) | 12 (60.0%) | 44 (73.3%) |
| **Subjects with solicited local VAAE** |  |  |  |  |
| Site pain | 16 (80.0%) | 12 (60.0%) | 9 (45.0%) | 37 (61.7%) |
| Swelling | 1 (5.0%) | 1 (5.0%) | 3 (15.0%) | 5 (8.3%) |
| Local heat | 2 (10.0%) | 2 (10.0%) | 1 (5.0%) | 5 (8.3%) |
| Redness | 4 (20.0%) | 2 (10.0%) | 2 (10.0%) | 8 (13.3%) |
| Induration | 1 (5.0%) | 0 (0.0%) | 0 (0.0%) | 1 (1.7%) |
| **Subjects with solicited systemic VAAE** |  |  |  |  |
| General malaise | 3 (15.0%) | 1 (5.0%) | 0 (0.0%) | 4 (6.7%) |
| Fever | 0 (0.0%) | 1 (5.0%) | 0 (0.0%) | 1 (1.7%) |
| **Solicited VAAE Total** | 50 (86.2%) | 28 (82.4%) | 21 (80.8%) | 99 (83.9%) |
| **Number of solicited VAAE per subject** |  |  |  |  |
| Average (SD) | 2.5 ± 2.1 | 1.4 ± 1.5 | 1.0 ± 1.5 | 1.6 ± 1.8 |
| Median (IQR) | 2.0 ± 2.8 | 1.0 ± 0.5 | 1.0 ± 1.8 | 1.0 ± 3.0 |
| Range | 0-8 | 0-5 | 0-6 | 0-8 |
| **Unsolicited VAAE Total** | 8 (13.8%) | 6 (17.6%) | 5 (19.2%) | 19 (16.1%) |
| **Number of unsolicited VAAE per subject** |  |  |  |  |
| Average (SD) | 0.4 ± 0.7 | 0.3 ± 0.6 | 0.2 ± 0.4 | 0.3 ± 1.8 |
| Median (IQR) | 0.0 ± 1.0 | 0.0 ± 0.8 | 0.0 ± 0.8 | 0.0 ± 1.0 |
| Range | 0-2 | 0-2 | 0-1 | 0-2 |

FINLAY-FR-1=dimeric-Receptor Binding Domain (d-RBD, 50 mcg) and outer membrane vesicles of *Neisseria meningitidis* group B (20 mcg) in aluminium hydroxide gel. FINLAY-FR-1-50= d-RBD (50 mcg) in aluminium hydroxide gel. FINLAY-FR-1-25= d-RBD (25 mcg) in aluminium hydroxide gel.
Data are n (%) unless otherwise specified. VAAE=Vaccine-Associated Adverse Event. Average (SD)=Average ± Standard Deviation. Median (IQR)=Median ± Interquartile Range.

In the group vaccinated with FINLAY-FR-1, the median value of solicited (2.0) vaccine-associated adverse events were higher than in the other two groups, although without statistically significant differences (Table 3). No significant changes were detected in laboratory tests after vaccination.

Very low antibody production was detected after the first dose in all groups. Anti-RBD seroconversion rates progressively increased according to the number of vaccine dose applied to each subject. After the third dose, seroconversion was 100%, 94.4% and 94.7% for the FINLAY-FR-1, FINLAY-FR-1A-50 and FINLAY-FR-1A-25 groups respectively. All formulations induced a very high increase in anti-RBD antibodies, with median values higher than those for baseline antibodies, those after the second dose (p<0.05) and those for the CCSP (Table 4).

**Table 4:**
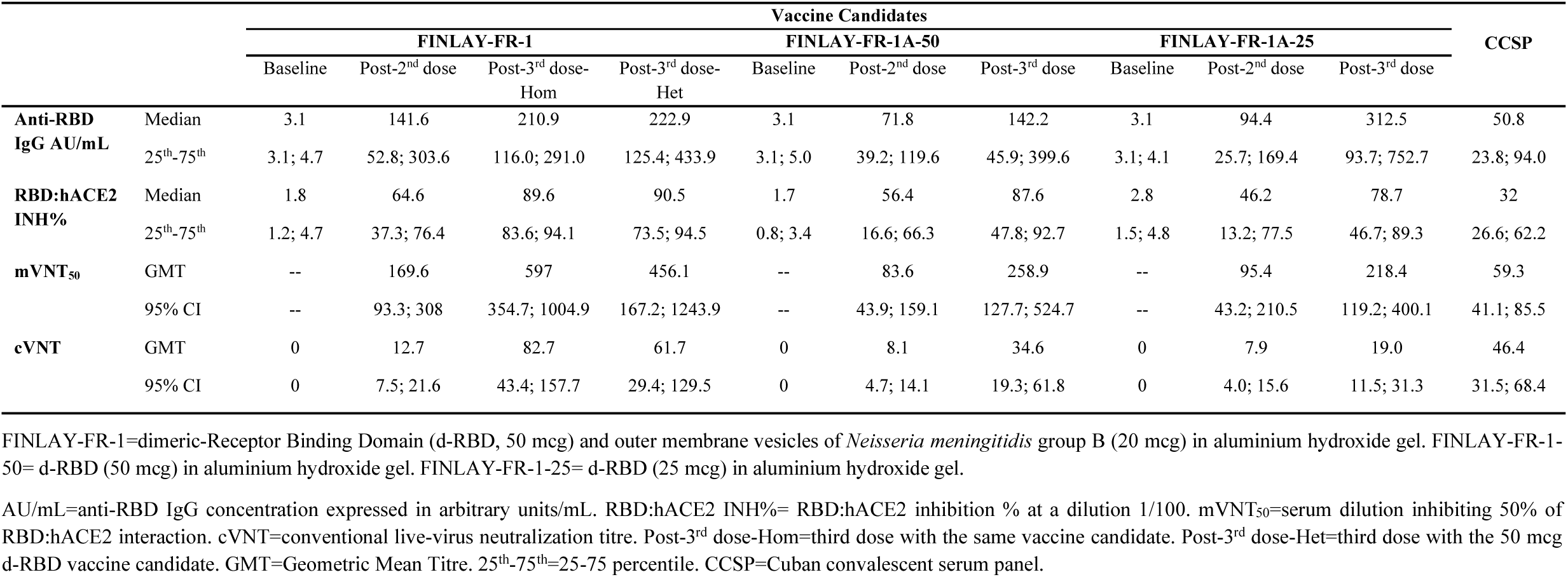
Humoral immune response induced by d-RBD vaccine candidates.

The inhibition of the RBD:hACE2 interaction increased notably after the second dose. A significant impact of the third dose was detected in the three groups (p<0.0001), with median values higher than those after the second dose and those for the CCSP (Table 4). All volunteers vaccinated with the complete FINLAY-FR-1 schedules achieved an RBD:hACE2 inhibition greater than the median value for CCSP.

The half-maximal molecular neutralization test (mVNT50) also showed a significant increase in functional antibody levels after the third dose (p<0.001), that was higher in the group vaccinated with FINLAY-FR-1 (p=0.028) (Table 4). All volunteers vaccinated with the FINLAY-FR-1 homologous schedule, and 90% of the volunteers with heterologous schedule achieved titres higher than the GMT of 68 sera included in the CCSP.

Live-virus neutralizing antibodies were detected after the second dose in 85%, 70% and 60% of the subjects vaccinated with FINLAY-FR-1, FINLAY-FR-1A-50 and FINLAY-FR-1A-25 respectively; increasing at 100%, 100% and 94.7% after the third dose. In FINLAY-FR-1 and FINLAY-FR-1A-50 groups, significant increases in GMT were detected with respect to the second dose (p<0.0001) (Table 4).

The GMT of live-virus neutralizing antibodies after the third dose increased significantly in the group vaccinated with both FINLAY-FR-1 schedules with respect to the other formulations (p=0.002) and to the CCSP (Table 4, Figure 2). Most subjects achieved titres higher than the GMT of CCSP: 90% with the homologous and 70% with the heterologous schedule. The FINLAY-FR-1A-50 group achieved higher GMT than the FINLAY-FR-1A-25 group after the third dose (p=0.002).

**Fig. 2.**
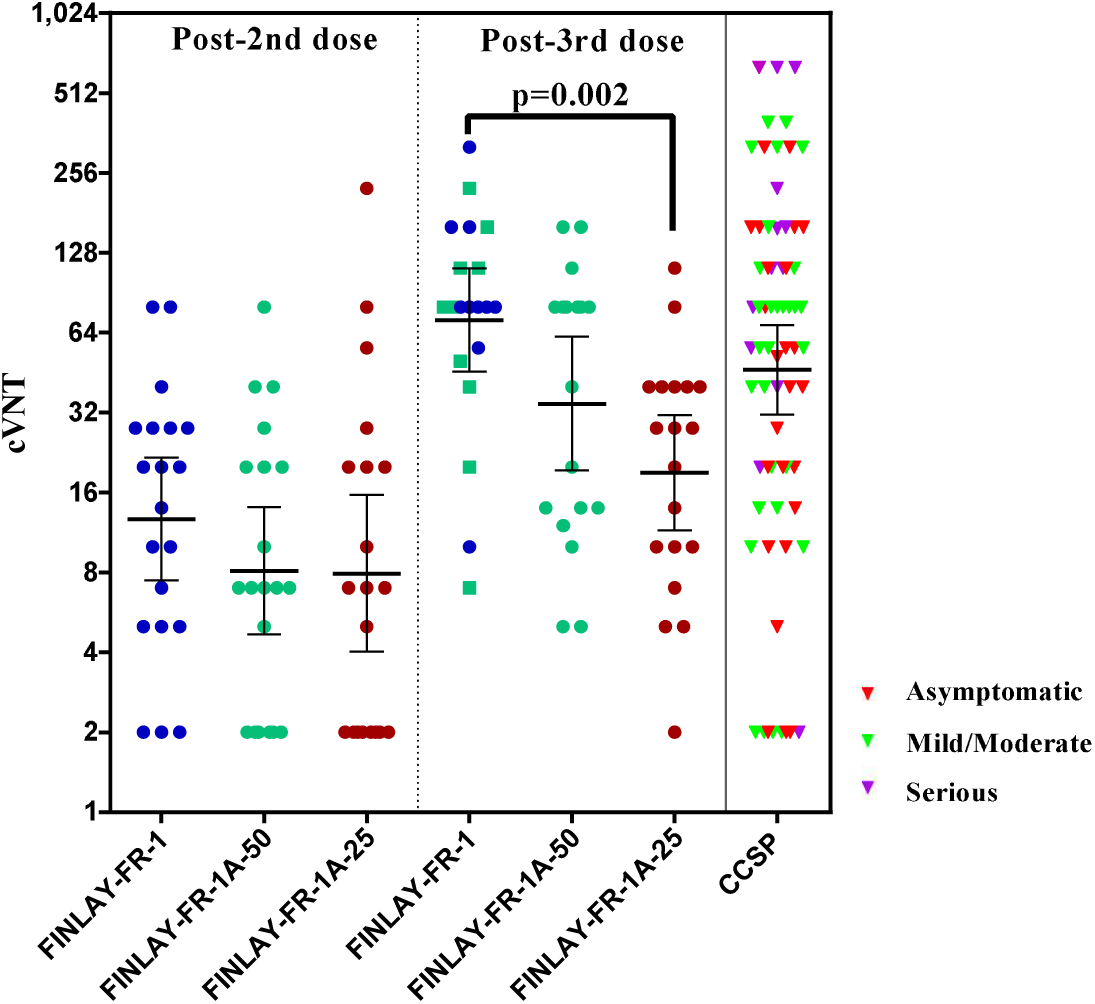
Conventional live-virus neutralization titres (cVNT) after second and third doses with the dimeric-Receptor Binding Domain (d-RBD) vaccine candidates: FINLAY-FR-1=dimeric-Receptor Binding Domain (d-RBD, 50 mcg) and outer membrane vesicles of *Neisseria meningitidis* group B (20 mcg) in aluminium hydroxide gel. FINLAY-FR-1-50= d-RBD (50 mcg) in aluminium hydroxide gel. FINLAY-FR-1-25= d-RBD (25 mcg) in aluminium hydroxide gel.

For all immunological endpoints analysed, no differences were found between FINLAY-FR-1 homologous and heterologous schedules. The 95% confidence intervals or 25-75 percentile ranges overlap, suggesting similarity in the immune response (Table 4).

FINLAY-FR-1 post-third dose=blue circles represent subjects with homologous schedules; green squares represent subjects with heterologous schedules. CCSP=Cuban convalescent serum panel.

## 4. Discussion

New generation vaccines, based on mRNA and viral vector vaccines are highly immunogenic [22-25]; however, there are concerns regarding their safety [9,10,22]. Vaccines based on conventional technologies, such as inactivated vaccines, are less immunogenic [10,22,26] and vaccines based on recombinant proteins, especially RBD, are probably less immunogenic, but cause fewer adverse reactions than new generation vaccines [10,22,27].

To improve the immunogenicity of subunit vaccines, new adjuvants are being used. These adjuvants activate antigen-presenting cells (APCs), which are necessary to stimulate T cells [22,27]. APCs are activated when they recognize microbe-associated molecular patterns (MAMPs) through pattern-recognition receptors, such as toll-like receptors (TLRs), C-type lectin-like receptors, and cytoplasmic receptors [13,28].

Some adjuvants, i.e. bacterial OMVs, can facilitate the antigen capture by APCs, especially dendritic cells, at the inoculation site and antigen delivery to the regional lymph nodes (signal 1 to T cell activation); antigen presentation to helper T cells and co-stimulation (signal 2); and immune polarization by cytokines (signal 3) [14,28].

The immunogenicity of our d-RBD vaccine candidates progressively increased with the number of vaccine doses. Also, it markedly improved when OMVs were present. OMVs from *Neisseria meningitidis* group B are an ideal adjuvant, suitable as both a delivery system, and a potent enhancer of the immune response [14,15]. OMVs are one of the components of the VA-MENGOC-BC^®^ vaccine. This is a bivalent meningococcal vaccine based on OMVs from group B and group C capsular polysaccharide. It has been successfully used in Cuba and other countries since 1989 to control epidemic meningococcal disease, with an excellent safety profile [29,30].

*Neisseria meningitidis* OMVs have also been used as adjuvant -named AFPL1-in several vaccine candidates. This adjuvant is a complex nano-structure that contains native lipopolysaccharide (LPS), PorB, and other MAMPs recognized by TLR-4, TLR-2 and TLR-9 receptors on APCs [14,29]. OMVs stimulate the production of IFNγ, IL-2, IL-12, pro-inflammatory cytokines (TNFα, IL-1β, IL-8) and chemokines (MIP1-α, MIP1-β). In addition, OMVs stimulate specific CD4+ helper T cells and CD8+ cytotoxic T cells, as well as the innate immunity. This adjuvant polarizes T helper responses to a Th1 pattern [14,15].

In short, OMVs activate the APCs, resulting in the activation of CD4+ helper T cells, followed by the activation of B cells and the production of specific neutralizing antibodies.

In this study, the efficacy of anti-RBD antibodies to block the interaction between recombinant RBD and hACE2 was evaluated in an inhibitory ELISA. All subjects vaccinated with FINLAY-FR-1 following the homologous or heterologous schedules developed a higher inhibition of the RBD:hACE2 interaction, greater than that of CCSP. According to the *in-vitro* molecular virus neutralization titres (mVNT_50_) and the live-virus neutralization test, —the gold standard to evaluate neutralizing antibodies against SARS-CoV-2—, most of the subjects vaccinated with both FINLAY-FR-1 schedules also achieved higher titres than those of CCSP.

This study demonstrated the adjuvant role of OMVs; functional antibodies, especially live-virus neutralizing antibodies markedly increased in this formulation. The neutralizing antibody titres of FINLAY-FR-1 were higher than those reported by inactivated virus vaccines [10,22,26] and some adenovirus vector vaccines, although lower than COVID-19 vaccines based on mRNA technology [22-25]. Additionally, OMVs stimulate the innate immunity that could help fight SARS-CoV-2.

The LPS component of OMVs slightly increased the reactogenicity of the FINLAY-FR-1 vaccine candidate; this reactogenicity is not higher than that reported for subunit vaccines with new adjuvants or those based on new technologies [9,22,27].

The first dose of a protein subunit vaccine, such as ours, triggers the primary immune response, stimulating naïve lymphocytes. Re-exposure to the same antigen induces a secondary immune response, qualitatively different, strong and rapid, due to activation of memory cells, responsible for the large increase of IgG anti-RBD and neutralizing antibodies.

Two doses elicit a secondary immune response; a third dose still improved the immune response appreciably, especially for FINLAY-FR-1 vaccine candidate. The immune response induced by the heterologous schedule with FINLAY-FR-1A (50 mcg) as a third dose was similar to the homologous schedule. As both responses are similar, the heterologous schedule is recommended, being the third shot (FINLAY-FR-1A—free of OMVs— instead of FINLAY-FR-1) less reactogenic.

The FINLAY-FR-1A vaccine candidate can be used as a booster for persons immunized with FINLAY-FR-1, immunized with other vaccines (due to the emergence of variants of concern, booster doses are being worldwide considered for other vaccines), and as a trigger of natural immunity in COVID-19 convalescents (a clinical trial, https://rpcec.sld.cu/en/trials/RPCEC00000349-En, has been accomplished, and a phase II clinical trial, https://rpcec.sld.cu/en/trials/RPCEC00000366-En, is ongoing).

Due to phase I results, regulatory authorities granted authorization for phase II clinical trial of the FINLAY-FR-1 vaccine candidate (https://rpcec.sld.cu/en/trials/RPCEC00000385-En)

## Data Availability

Information is available at the Cuban Public Registry of Clinical Trials, included in WHO International Clinical Trials Registry Platform (Soberana 01A, RPCEC https://rpcec.sld.cu/en/trials/RPCEC00000338-En). Other supporting clinical data and documents, including immunological individual data, will be available after publication of this article through direct request to: or.

## Acknowledgements

We thank the individuals enrolled in the study for their generosity, Dr Lila Castellanos-Serra for reviewing the manuscript, and the Soberana 01A Clinical Trial Team. Also, to the Ministry of Science, Technology and the Environment of Cuba for the partial funding of the Clinical Trial.

## Contributor Roles

SPR, MCRG and ROA are joint first authors. MCRG, BPM, YVB, DGR, ROA, and VVB conceived the study, designed the trial, the study protocol, and were involved in data analysis and interpretation. YCR, MMP, MAR, YCE and DGR supervised and monitored the trial. SPR, CAGD, ADM, MTR, AMT, JRM, MCRG, YCR, and BPMG were responsible for the site work including the recruitment and data collection. They contributed to data analysis and interpretation. LRN, BSR, RPN, THG, ASB, MDE, GBB, FPE and MTPG carried out immunological experiments and the analysis of results. RGM and CVS were involved in data curation and statistical analysis of data. GWC supplied resources. ROA and VVB wrote the manuscript, and all authors provided paper feedback.

## Data Sharing Statement

Information is available at the Cuban Public Registry of Clinical Trials, included in the WHO International Clinical Trials Registry Platform (Soberana 01A, RPCEC https://rpcec.sld.cu/en/trials/RPCEC00000338-En). Other supporting clinical data and documents, including immunological individual data, will be available after publication of this article through direct request to: or.

